# Ultra-sensitive Mapping of Cell-free DNA-derived Methylomes Reveals Brain-specific Epigenetic Alterations Implicated in Alzheimer’s Disease

**DOI:** 10.64898/2026.10.24.26366278

**Authors:** Xiaolong Cui, Alana V. Beadell, Shinya Tasaki, Ana W. Capuano, Zhou Zhang, Lu Gao, Brian C.-H. Chiu, Chuan He, David A. Bennett, Zoe Arvanitakis, Wei Zhang

## Abstract

DNA cytosine modifications play a crucial role in gene regulation that underlies the pathogenesis of complex diseases such as Alzheimer’s disease (AD). Using LABS (<u>L</u>inear <u>A</u>mplification-based <u>B</u>isulfite <u>S</u>equencing), an ultra-sensitive linear amplification-based low-input whole-genome methylation profiling method, we demonstrated the feasibility of detecting altered methylation features in serum-derived cell-free DNA (cfDNA) between a set of age- and sex-matched pathologically-defined participants with AD and non-AD controls, from the Rush Memory and Aging Project. These blood methylation changes were further linked to brain-specific epigenetic alterations implicated in AD. These findings support cfDNA methylome profiling as a promising non-invasive tool for AD biomarker discovery and research.

## Main

Alzheimer’s disease (AD) is a complex neurodegenerative disorder characterized by the progressive accumulation of amyloid-beta plaques and tau tangles^1,2^. Although the mechanism underlying AD pathogenesis is still not fully understood, previous studies have highlighted epigenomic modifications as a critical player during its development^3–9^. For instance, elevated histone acetylation H3K27ac/H3K9ac^3^ and altered DNA methylation^9^ have been linked to transcriptomic dysregulation in AD, indicating their roles as crucial gene regulators in AD biology. Importantly, these epigenetic alterations may also be detectable in circulating cell-free DNA (cfDNA), offering opportunities for minimally invasive biomarker development. Consistent with this concept, our recent study using a pull down-based technique (i.e., but without base resolution) showed biomarker potential of epigenetic features, specifically 5hmC detected in cfDNA for distinguishing AD from controls^10^. Blood-derived cfDNA holds considerable promise as a non-invasive source of epigenetic biomarkers for AD and multi-disease management^11–13^, yet this potential remains largely unexplored. Progress has been hampered by the limitations of both blood-derived cfDNA (e.g., often in the amount of a few nanograms from a blood draw) and enabling technologies (e.g., not base resolution, low genome coverage, high requirement of DNA materials, and high GC content bias). Here, we applied a novel technology called Linear Amplification-based Bisulfite Sequencing (LABS)^14^, an ultra-sensitive technique designed to overcome major challenges of existing techniques, to generate genome-wide DNA methylation profiles using serum-derived cfDNA samples collected from AD participants and age- and sex-matched non-AD individuals (controls). We sought to demonstrate the feasibility of detecting AD-relevant, brain-specific DNA methylation alterations at base resolution and exploring relation of blood to brain data, with the goal of laying foundation for future biomarker development in AD.

Specifically, we applied the LABS technique on an average of ∼1 ng cfDNA extracted from each serum sample^10,15^. A total of 32 LABS libraries were constructed (AD: n = 17; non-AD, n = 15) (**Fig. 1a, Table S1**). An average of 290 million paired-end reads were generated for each library, covering around 39.5 million CpG sites per sample. No obvious batch effect was detected in the genome-wide methylation data (**Extended Data Fig. 1a**). We observed classical metagene distribution in both sample groups, with low methylation levels at transcription start site (TSS) and transcription terminate site (TTS), and high methylation levels in gene bodies (**Extended Data Fig. 1b**). The linear amplification feature of LABS enables methylation quantification at CpG sites across the genome, even at loci with extreme GC percentages (**Extended Data Fig. 1c**). As a result, CpGs on repeat elements were well captured, with SINEs (short interspersed nuclear elements) representing the most profiled CpGs, followed by introns and intergenic regions (**Extended Data Fig. 1d**). In summary, we generated high-quality methylomes from cfDNA samples with single-base resolution from age- and sex-matched AD cases and non-AD controls.

**Figure 1.**
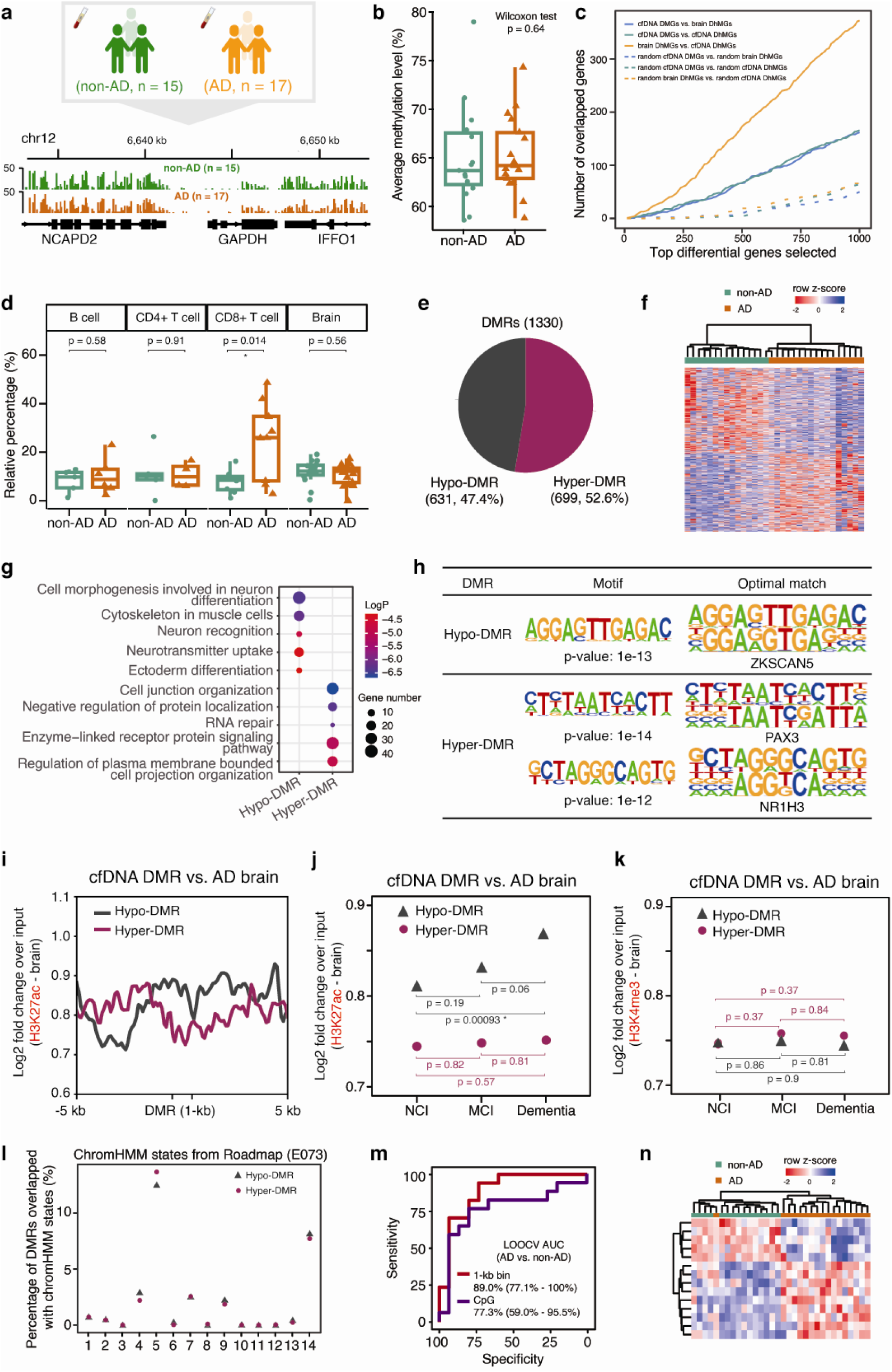
CfDNA methylomes reveal brain-specific epigenetic alterations. **a.** LABS data were obtained on serum-derived cfDNA samples collected from 32 study participants. Lower panel shows genome browser view of methylation levels around housekeeping gene *GAPDH*. **b.** Average methylation levels for all CpGs in each sample showing no significant differences between AD and non-AD groups. **c.** Differentially methylated genes from cfDNA overlapping with differentially hydroxymethylated genes from both cfDNA and brain. Dashed lines show number of overlapped genes from random sampling. **d.** Relative percentages of cell type-derived cfDNA fragments based on cellular deconvolution. **e.** Pie chart showing the number of hyper-methylated DMRs (hyper-DMR) and hypo-methylated DMRs (hypo-DMR). **f.** Heatmap showing the clustering pattern based on DMRs between AD and non-AD individuals. **g.** Functional enrichment plot for gene bodies holding DMRs. **h.** Motif analysis results for all DMRs, showing both de novo motifs and optimal match of known transcription factor binding motifs. Motif analysis was performed using Homer. **i.** Brain H3K27ac modification levels across DMRs from cfDNA methylomes. **j.** Average brain H3K27ac modification levels across DMRs from cfDNA methylomes along with different dementia levels. NCI, no cognitive impairment; MCI, mild cognitive impairment. **k.** Average brain H3K4me3 modification levels across DMRs from cfDNA methylomes along with different dementia levels. **l.** Percentages of DMRs overlapping with different chromHMM states. Brain prefrontal cortex chromHMM states were obtained from Roadmap Epigenomics Project (Epigenome ID: E073). **m.** The ROC plot showing the performance of 1-kb bin-based model and CpG-based model for AD. **n**. Hierarchical clustering of all samples based on 14 1-kb bins selected through elastic net regression.

Globally, DNA methylation levels, as represented by the average methylated levels, showed no significant differences between AD participants and controls (**Fig. 1b**). Next, to evaluate the feasibility of detecting cell/tissue sources for cfDNA profiles, we performed simulation analysis on top differentially methylated genes compared to top differentially hydroxymethylated genes from both cfDNA and brain samples^10^. More overlapped genes were observed, compared to random sampling of all gene sets, suggesting significant cfDNA contribution from the brain tissue (**Fig. 1c**). Moreover, we performed cellular deconvolution of the LABS data based on published reference methylomes that cover 13 cell and tissue types, following our previous study^14^. As expected for blood-derived cfDNA, neutrophils were found to contribute the most to the cfDNA pool (**Extended Data Fig. 2**). Notably, we indeed observed considerable brain-derived signals in cfDNA methylomes, suggesting the feasibility of using cfDNA as a surrogate for epigenetic changes in AD brain. In addition, a significant change in the relative proportions of CD8+ T cell-derived sources weas observed between AD and controls (**Fig. 1d**), consistent with the involvement of the adaptive immune system in AD^16^.

To systematically characterize altered DNA methylation associated with AD samples, we performed genome-wide differential analysis on the unbiased 1-kb sliding windows, in models adjusting for sex. A total of 1330 DMRs (differentially methylated regions) were detected (q-value < 0.05, methylation difference > 0.2, **Fig. 1e, Table S2**), with 631 DMRs showing hypomethylation in AD (hypo-DMRs), and 699 hypermethylated in AD (hyper-DMRs). Hierarchical clustering based on the 1330 DMRs readily separated the two sample groups, indicating the feasibility of identifying non-invasive methylation biomarkers to distinguish AD from controls (**Fig. 1f**).

We further annotated the DMRs to different genomic elements (**Extended Data Fig. 3a, 3b**). As expected, most DMRs were located in intergenic regions and introns, consistent with the total lengths of these genomic regions and the distributions of genome-wide CpGs. Additionally, we also observed considerable DMRs located in repeat elements, such as LINEs (long interspersed nuclear elements), LTRs (long terminal repeats), and SINEs, suggesting universal changes across different genomic elements in AD. To provide biological insights into those AD-associated DMRs, we performed functional enrichment analysis for the host genes in DMRs (**Fig. 1g**). Intriguingly, brain-related functions were found to be highly enriched (FDR < 0.05), suggesting that cfDNA profiles could capture AD brain-relevant pathways and biological processes. Notably, the hypo-DMRs were enriched for functional terms related to brain development, such as cell morphogenesis involved in neuron differentiation, neuron recognition, and ectoderm differentiation, while the hyper-DMRs were enriched for functional terms related to relatively fundamental pathways, such as RNA repair, and enzyme-linked receptor protein signaling pathway. In line with our findings, recent single-cell ATAC-seq data from hippocampi also suggested that neuronal developmental genes were activated through elevated chromatin accessibility^17^. Interestingly, using motif analysis^18^, we found highly significant motifs for both hypo-DMRs and hyper-DMRs (**Fig. 1h**), while the optimal matched motif analysis showed transcription factors (i.e., *ZKSCAN5*) known to play roles in AD biology^19–21^.

We further examined whether cfDNA methylomes were also implicated in brain-derived epigenomic changes associated with AD. We first compared our cfDNA DMRs with published brain DMRs^9^, but found no overlap, possibly due to the limited genomic coverage of Illumina HumanMethylation 450k beadchip used in that study. Previous research has also found elevated histone acetylation in AD brains compared to non-AD controls^3^. Therefore, we obtained brain-derived H3K27ac ChIP-Seq datasets from the ENCODE Project (n = 5) and summarized the H3K27ac levels across cfDNA-based DMRs (**Fig. 1i**). Indeed, high H3K27ac levels were observed for hypo-DMRs, while low H3K27ac levels were observed for hyper-DMRs, suggesting that cfDNA-based DMRs could potentially reflect brain-derived enhancers. Moreover, cognitive impairment levels were found to be associated with a trend of higher H3K27ac levels for hypo-DMRs, while no trend of changes was observed for hyper-DMRs (**Fig. 1j**). In contrast, there were no similar trends between cfDNA-based DMRs and the two brain-derived histone methylations: H3K27me3 and H3K4me3 (**Fig. 1k, Extended Data Fig. 4a, 4b, 4c**).

We asked whether we could observe the epigenome erosion of AD^22^ from blood cfDNA methylomes. We obtained prefrontal cortex chromHMM states from Roadmap Epigenomics Project and compared to our DMRs from cfDNA (**Fig. 1l**). Intriguingly, hyper-DMRs have larger active chromatin state (chromHMM state 5) compared to hypo-DMRs, while hypo-DMRs have larger repressed chromatin state (chromHMM state 14), suggesting repression of active chromatin regions and activation of repressed chromatin regions. Therefore, cfDNA methylomes reflected brain-derived epigenomic modifications, supporting that blood cfDNA can be a surrogate for AD brain and the LABS as a powerful tool that can facilitate AD research.

We sought to explore the feasibility of summarizing genome-wide LABS data into an epigenetic model for distinguishing AD from controls that could be further developed and validated. Limited by the relatively small sample size, we used the leave-one-out-cross-validation (LOOCV) method to demonstrate modeling performance using elastic net regularization and multivariable logistic regression models, following our previous studies^7^. Specifically, a model based on selected 1-kb bins achieved an average AUC (area under the curve) of 89.0% and 95% confidence intervals (CIs) of 77.1% to 100%. Given the possibility of obtaining CpG-level modifications, we also explored the feasibility of modeling based on single CpGs using DMRs as candidates, from which the final model showed an average AUC of 77.3% and 95% CIs of 59.0% to 95.5% (**Fig. 1m**). Additionally, 14 1-kb bins with > 70% frequency in our LOOCV feature selection process can readily separate AD from control by hierarchical clustering (**Fig. 1n**). Our machine learning-based modeling demonstrated the feasibility of developing non-invasive biomarkers using the LABS in cfDNA for AD in future studies based on larger sample sizes.

In conclusion, in the current study, we used an ultra-sensitive genome-wide methylome profiling technique developed by our team to test circulating cfDNA samples from AD research participants and matched non-AD individuals and revealed cfDNA methylomes associated with AD brains. The feasibility of detecting AD brain-associated epigenomic changes from circulating cfDNA indicated broad opportunities of using clinically convenient blood-derived cfDNA as a surrogate for brain tissue to study genomic and epigenomic etiologies in AD. Moreover, we provided here a proof-of-concept analysis that non-invasive biomarkers for AD detection could be achieved by LABS in cfDNA assay as well. Future investigations utilizing larger and other cohorts, are warranted to facilitate a more systematic and comprehensive characterization of the AD epigenome and biomarker discovery. Findings from the genome-wide LABS assay in cfDNA could also provide candidates for site-specific and less expensive detection methods, such as our team’s mass spectrum-based approach – LAMP-MS (Linear Amplification coupled with Matrix-Assisted Laser Desorption/Ionization Time-of-Flight

Mass Spectrometry on silicon chips)^23^, with the ultimate goal of developing clinically convenient, robust and affordable tests for precision medicine.

## Methods

### Study population and clinical samples

Details about selection of study participants have been described in a previous study. Briefly, a subset of deceased and autopsied persons were selected from the Rush Memory and Aging Project (MAP), an epidemiologic longitudinal, clinical-pathologic study, enrolling mostly non-Latino White participants (70-95 years old) since 1997.^15^ Participants were matched using a Mahalanobis distance-based algorithm, such that 4 groups of 20 selected participants with a similar distribution of age at death (within 5 years) and sex were obtained.

### Preparation of LABS library and sequencing

Extracted cfDNA was ligated with T7 adaptor using KAPA Hyper Prep Kit (Roche Cat. No. KK8504). Briefly, DNA in nuclease-free H2O was mixed with end repair and A-tailing buffer and end repair and A-tailing enzyme mix and incubated at 20 °C 30 min and 65 °C 30 min. Next, the homemade T7 adaptor, H2O, ligation buffer, and DNA ligase were added and incubated at 20 °C for 4 h or 4 °C overnight. After post-ligation clean-up using 1.4X AMPure XP Beads, DNA was subjected to bisulfite treatment. Bisulfite-converted DNA was extended via using EpiMark Hot Start Taq DNA Polymerase (NEB, M0490S) with T7 Primer (AGCCCCGCGAAATTAATACGACTCACTATAGGG, IDT with HPLC purification). Qiagen protease (Qiagen, Cat. No. 19155) was added and incubated at 50 °C for 2 h, followed by heat inactivation at 75 °C for 30 min. The T7-tagged dsDNA fragments were used as a template to generate in vitro transcription via HiScribe™ T7 High Yield RNA Synthesis Kit (NEB, E2040S) and SUPERase•In™ RNase Inhibitor (Life Technologies, AM2694) and were added in the reaction. The reaction was incubated at 37 °C for 12–16 h.

After overnight T7 in vitro transcription, DNase I (NEB, M0303S) and digestion buffer were added to digest DNA templates at 37 °C for 10 min. Then, RNA transcripts were purified by RNA Clean and Concentrator kit (Zymo Research, R1013). RNA yield was quantified by Qubit 2.0 RNA HS Assay Kit (Life Technologies, Q32855). A maximum of 100 ng RNA was used for library construction (Roche, KAPA RNA HyperPrep kit, KK8541) following the manufacturer’s protocol. Libraries were sequenced at the University of Chicago Genomics Core Facility.

### Bioinformatic processing of LABS data

Sequencing adaptors and low-quality nucleotides were trimmed from raw sequencing reads by Trim_Galore and then aligned to the human reference genome (hg19) by Bismark according to a published processing procedure. Further duplication removal and methylation calling were performed by scripts of the Bismark package. The methylation proportion of each CpG (at least 3 ×) was determined using the methylKit R package. Besides CpG-level data, for analytical convenience, data was also summarized into unbiased, genome-wide 1-kb sliding windows for downstream analyses, represented by the weighted mean of CpGs for each bin. Differential methylation regions were detected by methylKit R package, with a cutoff of FDR < 0.05 and methylation differences larger than 20%. Metagene plots were generated by deepTools. GC content bias was assessed by Picard tools. IGV was used for the genome browser view. Assignment of CpGs to different genomic features (e.g., exons, introns) was done by Homer with a default priority list. Functional enrichment analysis was performed using Metascape online tool for genes with DMRs. Cellular deconvolution was calculated by CIBERSORT (https://cibersortx.stanford.edu/) tool.

### Integration with public omics datasets

ChIP-Seq datasets for histone modifications H3K27ac, H3K4me3, H3K27me3 from human brain samples with various levels of cognitive impairment (no cognitive impairment; mild cognitive impairment; dementia attributed to Alzheimer’s Disease) were downloaded from ENCODE project portal (Rush Alzheimer’s Disease Study). For each cognitive impairment level, we selected 5 samples. Genome-wide 5hmC profiles of the same study participants in our current study (n = 27) were previously obtained. The experimental protocol and bioinformatic processing were described in a previous publication.^10^ Average profiles for different omics datasets were generated using deepTools (https://deeptools.readthedocs.io/en/latest/).

### Machine learning-based feature selection and modeling

Due to the small sample sizes, leave-one-out-cross-validation was performed in R using *caret* package. Elastic net was used for feature selection, and logistic regression was used for modeling. The receiver operating characteristic curve was generated by *pROC* R package, and an AUC was computed with 95% CI to assess the performance of the model.

### Statistical analysis

All statistical analyses were conducted using the R Statistical Environment (v4.4.0). Unless otherwise stated, statistical significance between two groups was tested using Wilcoxon signed-rank tests.

## Data availability

The LABS data generated in this study is available from the corresponding authors upon request. ChIP-Seq datasets for histone modifications H3K27ac, H3K4me3, H3K27me3 from human brain samples are available at the ENCODE project portal for Rush Alzheimer’s Disease Study: https://www.encodeproject.org/brain-matrix/?type=Experiment&status=released&internal_tags=RushAD. Data from the Rush Alzheimer’s Disease Center, including demographic, cognitive, and other data about the Memory and Aging Project are available by submitting a data request at: www.radc.rush.edu; following a committee review for approval, and the requestor will need to sign a data use agreement.

## Data Availability

All data produced in the present study are available upon reasonable request to the authors

## Acknowledgements

We thank Dr. Pieter W. Faber and the staff at the Genomics Facility and Comprehensive Cancer Center sequencing facility at the University of Chicago for performing the NGS sequencing measurements. We thank support from the National Institute of Health (NIH) grants: R01 AG074549, R33 CA269100, R01 AG17917.

