## supplemental figures for "Ultra-sensitive Mapping of Cell-free DNA-derived Methylomes Reveals Brain-specific Epigenetic Alterations Implicated in Alzheimer’s Disease"

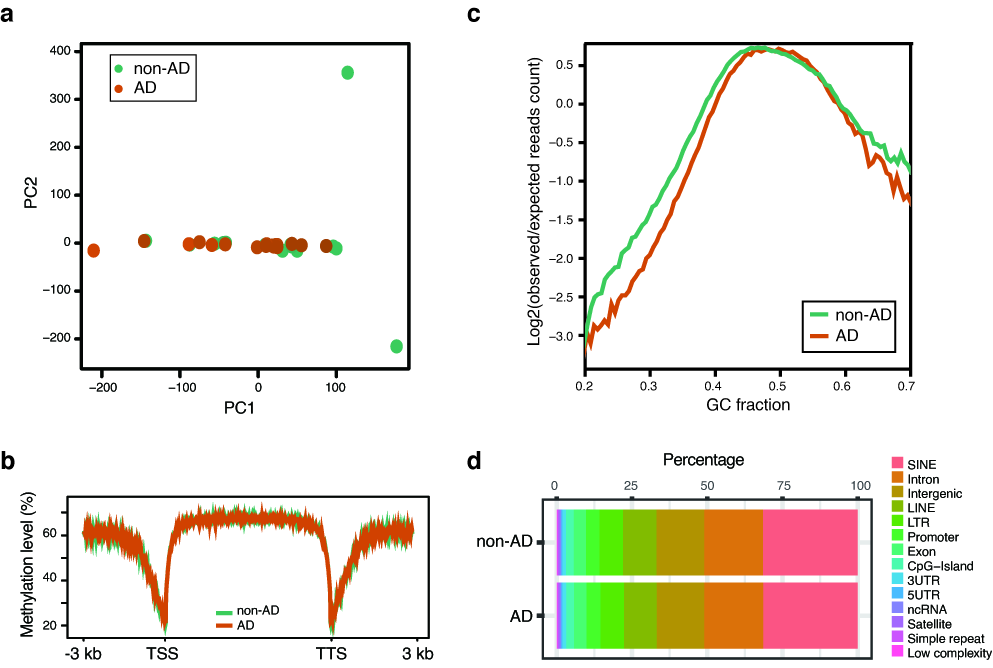

**Extended Data Figure 1. Overview of LABS data on circulating cfDNA. a.** PCA plot based on CpG methylation levels indicating no obvious systematic bias across the 32 cfDNA samples. **b.** Metagene plot showing methylation distribution in all profiled cfDNA samples. TSS, transcription start site; TTS, transcription terminate site. **c.** Line plot showing the reads coverage across genomic regions with different GC fractions. **d.** Bar plot showing distributions of LABS covered CpGs across the genome.

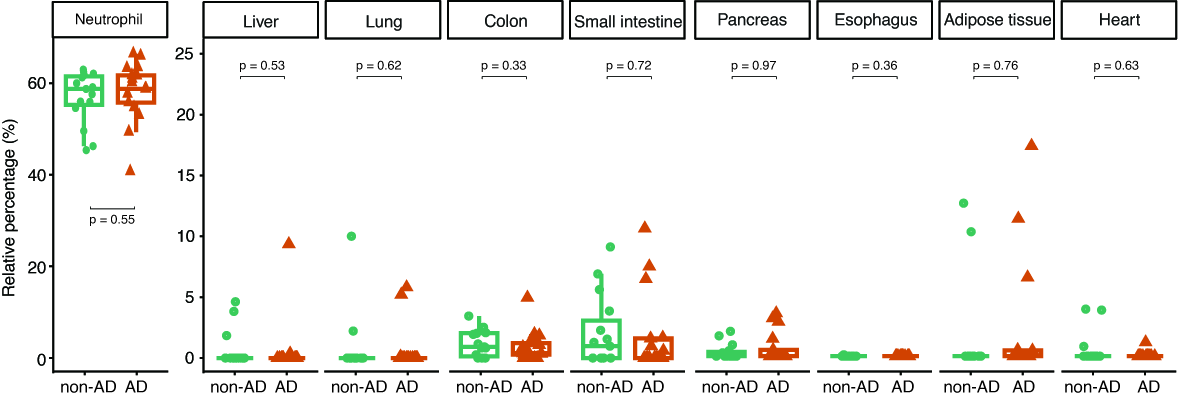

**Extended Data Figure 2. Relative percentages of cell type-derived cfDNA fragments based on cellular deconvolution.** Cellular deconvolution was performed by CIBERSORT.

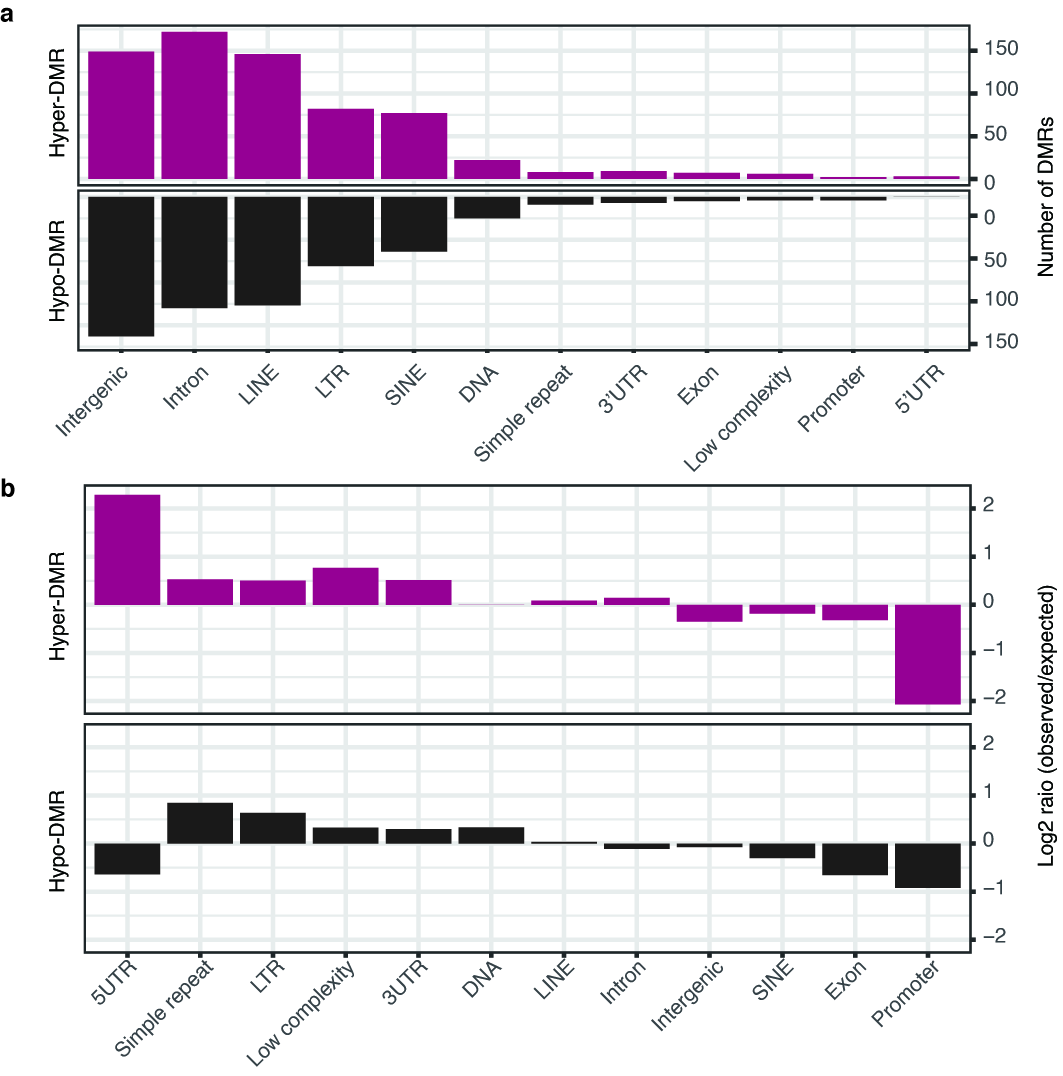

**Extended Data Figure 3. Characteristics of AD-specific DMRs from cfDNA methylomes. a.** Distributions of AD-specific DMRs across different genomic elements. **b.** Relative enrichment of AD-specific DMRs across different genomic elements.

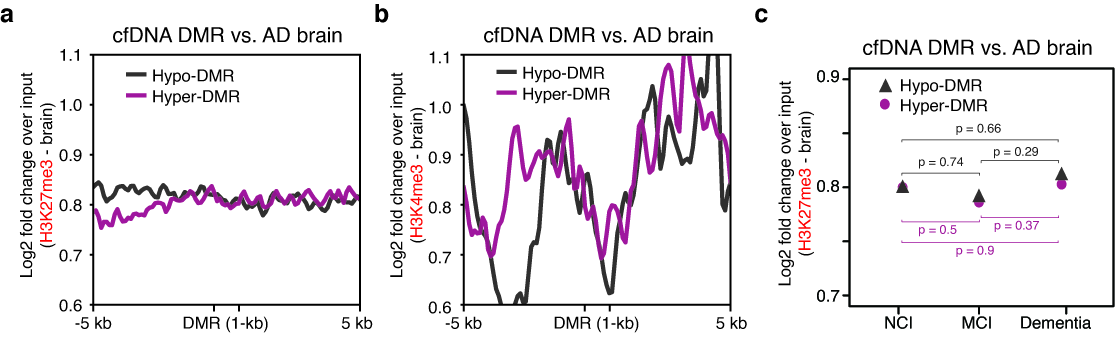

**Extended Data Figure 4. Histone modification levels in brain across cfDNA-based DMRs. a.** Brain H3K27me3 modification levels across DMRs from cfDNA methylomes. **b.** Brain H3K4me3 modification levels across DMRs from cfDNA methylomes. **c.** Average brain H3K27me3 modification levels across DMRs from cfDNA methylomes along with different dementia levels.

**Supplementary Table 1. Demographic information of all 32 study participants.**

**Supplementary Table 2. Differentially methylated regions generated from cfDNA methylomes.**
